# Neutralizing anti-DNase1L3 antibodies derive from autoreactive VH4-34^+^-B cells and associate with the interferon signature in SLE

**DOI:** 10.1101/2021.06.07.21258180

**Authors:** Yikai Yu, Eduardo Gomez-Bañuelos, Jessica Li, Kevin S. Cashman, Merlin Paz, Maria Isabel Trejo-Zambrano, Regina Bugrovsky, Youliang Wang, Asiya Seema Chida, Cheryl A. Sherman-Baust, Dylan P. Ferris, Daniel W. Goldman, Erika Darrah, Michelle Petri, Iñaki Sanz, Felipe Andrade

## Abstract

DNase1L3 deficiency is an inborn error of immunity that causes monogenic systemic lupus erythematosus (SLE) in humans. Here, we identified that one third of patients with sporadic SLE have antibodies to DNase1L3. Like DNase1L3 deficiency, we found that patients with anti-DNase1L3 antibodies have features associated with immune pathways activated by immunogenic self-DNA, including elevated antibodies to dsDNA and prominent expression of the interferon and myeloid/neutrophil signatures. Interestingly, 40-80% of anti-DNase1L3 antibodies in SLE serum contain the 9G4 idiotype, which is encoded by the autoreactive heavy-chain gene segment V_H_4-34. Sequence and functional analysis of four anti-DNase1L3 monoclonal antibodies generated from SLE patients experiencing disease-associated flares showed that these antibodies were derived from self-reactive 9G4^+^ switched memory B cells. These antibodies are highly enriched in somatic hypermutations, indicating that they originated from antigen-experienced cells, and have neutralizing activity against DNase1L3. Together, the data demonstrate that autoantibodies to DNase1L3 phenocopy pathogenic mechanisms associated with DNase1L3 deficiency. Moreover, the finding that autoreactive B cells bearing the 9G4 idiotype produce dominant serum autoantibodies, including antibodies to DNase1L3, underscores V_H_4-34^+^ B cells as sensible therapeutic targets for specific depletion of pathogenic B cells in SLE.

## INTRODUCTION

Systemic lupus erythematosus (SLE) is a complex multisystem disease characterized by the production of antibodies to a diverse number of autoantigens and immune-mediated tissue damage (1). While in most of the cases the etiology of SLE is unknown, there is a small proportion of patients in which the disease is the result of a monogenic inborn error (2). Human inborn errors of immunity are single gene defects that can be manifested by increased susceptibility to infections, autoimmunity, autoinflammation, allergy, and/or malignancy (3). Some inborn errors of immunity affecting soluble factors, such as cytokines, can also be phenocopied by the production of neutralizing autoantibodies against those proteins (4). In the case of monogenic causes of lupus and lupus-like syndromes, these are commonly associated with defects in genes encoding proteins involved in the clearance of nucleic acids, both intra and extracellularly (2). Among these defects, deficiency of DNase1L3 has been recently associated with the development of SLE (5, 6).

DNase1L3 is a member of the DNase1 family of DNA endonucleases. The enzyme is primarily secreted by myeloid cells (i.e. macrophages and dendritic cells) (7-9), and together with DNase1, is responsible for the DNase activity in circulation (10). Different to DNase1, however, DNase1L3 contains a positively charged C-terminal sequence that permits it to digest liposome-complexed DNA and displace DNA from bound histones (11, 12). Thus, DNase1L3 is more efficient than DNase1 in the inter-nucleosomal cleavage of nuclear DNA, suggesting that its major role is the digestion of chromatin from apoptotic and necrotic cells (11, 13). Null mutations and hypomorphic variants of DNase1L3 are linked to familial and sporadic SLE, respectively (5, 6). In addition, lupus-prone MRL and NZB/W F1 mice are deficient in DNase1L3 and the sole deficiency of this enzyme leads to lupus-like disease in mice (11, 14). Mechanistically, DNase1L3 decreases the availability of antigenic cell-free DNA by fragmenting DNA, reducing its exposure on apoptotic cell microparticles (11, 15). In the absence of DNase1L3 activity, extracellular self-DNA drives TLR-dependent IFN-I production and extrafollicular differentiation of antibody-forming cells, driving anti-dsDNA antibodies and SLE (16).

Here, we showed that a subset of patients with SLE develop antibodies to DNase1L3. Patients with anti-DNase1L3 antibodies are characterized by a significant increase in disease activity, elevated antibodies to diverse autoantigens (particularly dsDNA) and prominent transcriptional expression of the interferon and myeloid/neutrophil signatures. Through the analysis of SLE patient-derived monoclonal antibodies, we found that this new subset of autoantibodies is antigen-driven and derived from autoreactive V_H_4-34-expressing IgG switched memory B cells. Moreover, we also demonstrate that these autoantibodies have neutralizing activity to DNase1L3. Together, our findings shed light on the antigenic repertoire of pathogenic antibodies produced by inherently autoreactive V_H_4-34^+^ cells in SLE and identify anti-DNase1L3 antibodies as potential amplification drivers amenable to therapy in this disease.

## RESULTS

### DNase1L3 is a target of autoantibodies in SLE

To gain insights into the role of DNase1L3 in SLE pathogenesis, we studied the expression and activity of this enzyme in peripheral blood mononuclear cells (PBMC) and serum, respectively. Unexpectedly, we found that patients with SLE had a prominent and significant increase in the transcriptional expression of DNase1L3 in PBMCs compared to healthy controls (**Figure 1A**). To define the relationship between DNase1L3 expression and activity, DNase1L3-mediated chromatin degradation was determined in samples in which matched PBMC/sera were available. Different to DNase1, DNase1L3 has a unique capacity to degrade native chromatin and therefore, serum/plasma-mediated chromatin degradation has been used as an indirect method to quantify DNase1L3 activity in circulation (17, 18). However, DNase1 in serum degrades chromatin effectively in cooperation with serine proteases, such as plasmin or thrombin, which remove DNA-bound proteins (10). In addition, it is suspected that DNase1L3 in serum is sensitive to proteolysis or indirectly inhibited by proteolysis of DNA-bound structural proteins (10). Therefore, in the absence of protease inhibitors, chromatin degradation assays using serum cannot distinguish between the activities of DNase1 and DNase1L3 (10). Similarly, using plasma (regularly collected with heparin or calcium chelators) to study DNase1L3 activity has important limitations. Heparin displaces histones from chromatin, facilitating chromatin degradation by DNase1 (10). In addition, heparin inhibits DNase1L3 (10). Recalcification of EDTA or citrated plasma (required to activate DNase1L3) activates the coagulation pathway, which can enable chromatin breakdown by DNase1 and thrombin/plasmin. Thus, to avoid these numerous caveats, we used serum (instead of plasma) treated with aprotinin to inhibit serine proteases and chromatin degradation was measured using intact nuclei (10).

**Figure 1.**
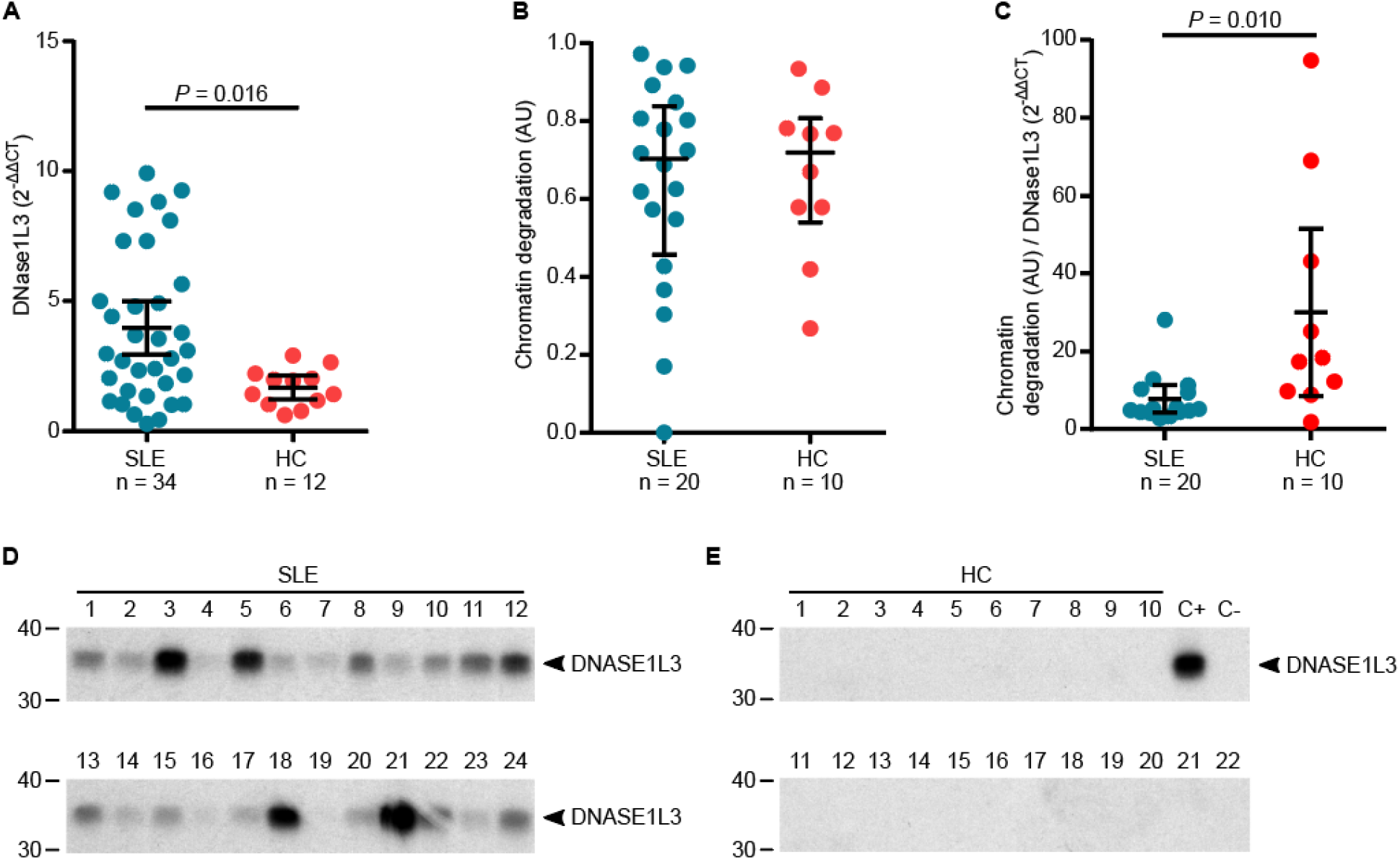
Patients with SLE have antibodies to DNase1L3. (**A** and **B**). DNase1L3 transcriptional expression in PBMCs (**A**) and activity in serum (**B**) were determined by qPCR and chromatin degradation assays (Scale 0 = no degradation, 1 = complete degradation), respectively. (**C**) Ratio of DNase1L3 serum activity per DNase1L3 expression 2^−ΔΔC^_T_ in PBMCs. Comparisons were done using Mann-Whitney’s U test. (**D** and **E**) Anti-DNase1L3 antibodies were determined in serum by immunoprecipitation of radiolabeled DNase1L3 in a convenience sample of SLE patients with active disease (n=24) (**D**), and 22 healthy controls (n=22) (**E**). In **E**, SLE sera #3 and #19 (from **D**) were used as positive (C+) and negative (C-) controls, respectively.

Interestingly, although SLE patients had higher levels of DNase1L3 transcriptional expression (**Figure 1A**), we found no significant differences when comparing DNase1L3 activity in control and SLE serum (**Figure 1B**). Indeed, there was a significant discrepancy between the expression and the activity of the enzyme in SLE compared to controls (**Figure 1C**). We hypothesized that in some patients, this paradox may result from compensatory mechanisms trying to overcome a potential inhibition of DNase1L3 in SLE. To explore this idea, we initially searched for antibodies to DNase1L3 in healthy control and SLE serum from a convenience sample of patients with active disease. Remarkably, almost every SLE serum tested showed some reactivity to DNase1L3 (**Figure 1D**), with clear variation among samples likely resulting from different anti-DNase1L3 antibody levels. In contrast, control serum showed no reactivity against this enzyme (**Figure 1E**).

### Anti-DNase1L3 antibodies are associated with clinical and immunological features of active SLE

To define the prevalence and clinical associations of antibodies to DNase1L3 in patients with SLE, we studied a prospective observational cohort for which extensive clinical and serologic data are available, as well as whole blood gene expression analysis (19). Demographic, clinical and laboratory features of the SLE cohort are summarized in **Supplemental Table 1**. Antibody reactivity to DNase1L3 was significantly increased in SLE compared to healthy controls (*P* < 0.001) (**Figure 2A** and **Supplemental Figure 1**). Using a cutoff of two standard deviations above the mean anti-DNase1L3 antibody level in healthy sera, 30% (48/158) of SLE patients *versus* 1.6% (1/62) of healthy controls were positive for anti-DNase1L3 antibodies (*P* < 0.0001). Antibodies to DNase1L3 were significantly associated with anemia, livedo, proteinuria, low complement and a broad range of autoantibodies, including anti-dsDNA, anti-cardiolipin, lupus anticoagulant, anti-β2-glycoprotein I, and anti-Ro52 antibodies (**Figure 2B** and **Supplemental Table 1**). SLE patients with anti-DNase1L3 antibodies also have features suggestive of a history of more severe disease, including clinical signs of secondary Cushing’s syndrome (i.e. Moon facies) and the use of cytotoxic treatment (**Figure 2B** and **Supplemental Table 1**).

**Figure 2.**
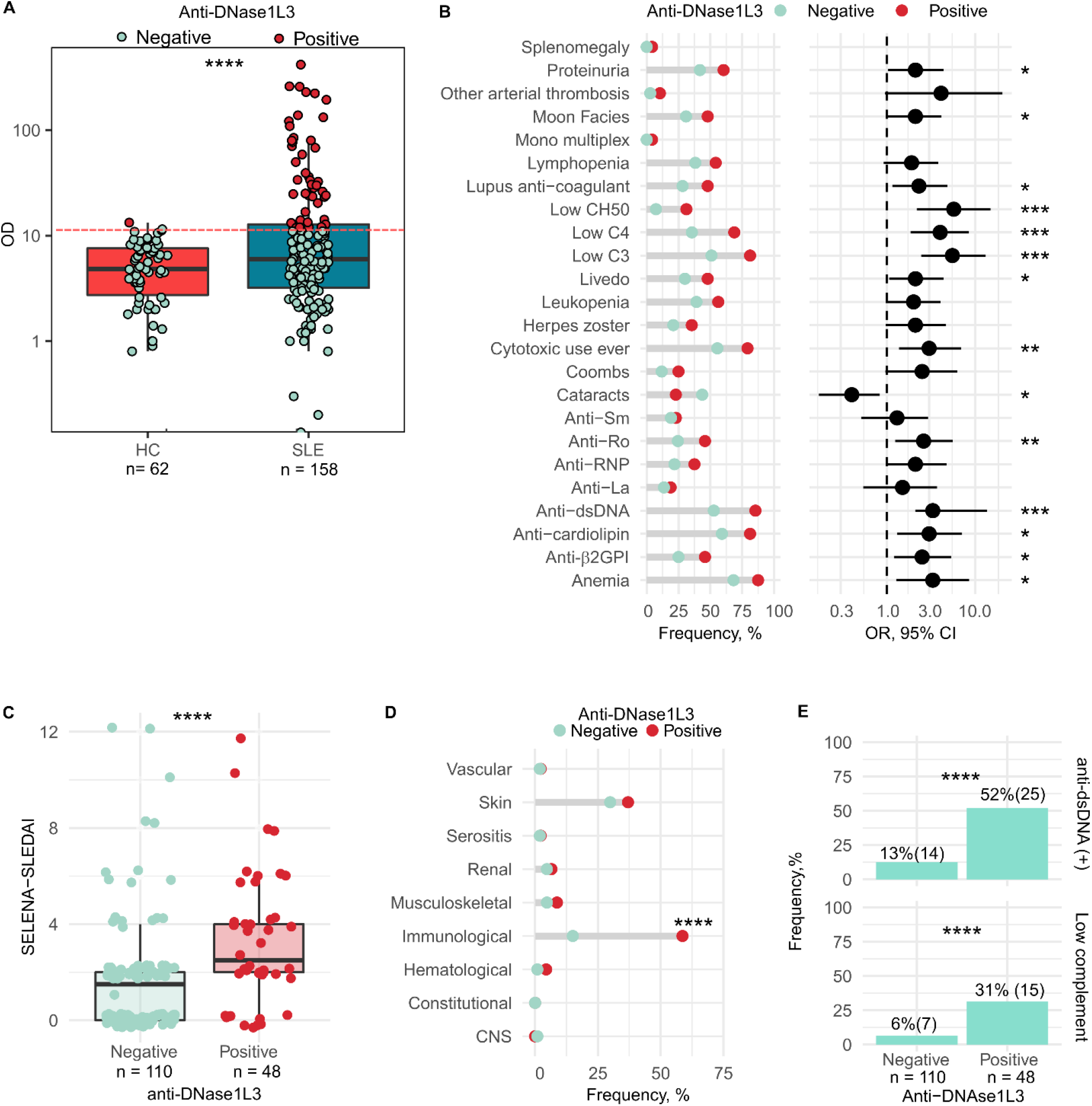
Prevalence and clinical associations of antibodies to DNase1L3 in a prospective observational cohort of patients with SLE. (**A**) Serum levels and positivity of anti-DNase1L3 antibodies in healthy controls (HC) and patients with SLE. Using a cutoff of two standard deviations above the mean anti-DNase1L3 antibody level in healthy sera, 1.6% (1/62) of healthy controls and 30% (48/158) of SLE patients were positive for anti-DNase1L3 antibodies. (**B**) Associations between anti-DNase1L3 antibodies and clinical/serologic features in SLE. Bars represent the frequency of clinical/serologic features according to anti-DNase1L3 antibody status (left) and their corresponding OR with 95% CI (right). (**C**) Safety of Estrogens in Lupus National Assessment study-SLE disease activity index (SELENA-SLEDAI) of SLE subjects, at time of visit, according to anti-DNase1L3 positivity. (**D**) Associations between individual SELENA-SLEDAI score items ‘at time of visit’ and anti-DNase1L3 antibodies. SELENA-SLEDAI items are represented as categorical variables. CNS: central nervous system. (**E**) Frequency of SLE subjects positive for anti-dsDNA antibodies (upper) and low complement ‘at time of visit’ according to anti-DNase1L3 status. Comparisons of continuous variables were done using Student’s τ test. Associations between categorical variables were performed with χ^2^ of Fisher’s exact tests accordingly. \*\*\*\**P* < 0.0001, \*\*\**P* < 0.001 and \*\**P* < 0.01.

At time of visit, anti-DNase1L3 antibodies were significantly associated with higher disease activity by SELENA-SLEDAI [median, (IQR), 1.8 (0-12) *vs*. 3.4 (0-12), *P* = 0.002] (**Figure 2C**), which was determined by the immunological domain (**Figure 2D**). Specifically, elevated anti-dsDNA [12.4 (7/113) *vs*. 55.6 (15/45), *P* < 0.0001] and low complement [6.2 (7/113) *vs*. 33.3 ((15/45), *P* < 0.0001] were more common with anti-DNase1L3 antibodies (**Figure 2E**). Further, at the visit, anti-DNase1L3 positive patients were more likely treated with prednisone and cytotoxic drugs (**Supplemental Figure 2A**) and received higher doses of prednisone *versus* anti-DNase1L3 negative patients [median (IQR), 5(0-6.88) *vs*. 0(0-4), *P* = 0.0001] (**Supplemental Figure 2B**).

In twenty-four SLE patients for whom longitudinal data were available, the levels of anti-DNase1L3 antibodies showed different patterns overtime. In most of the patients, the antibody status remained stable in serial samples (**Supplemental Figure 3 and 4**). However, in a subset of anti-DNase1L3 positive patients, antibody levels varied overtime and in some cases, there was a clear association between antibody levels and SLE disease activity (**Supplemental Figure 3**). The data, however, were limited and unable to define whether anti-DNase1L3 antibodies can predict future flares.

**Figure 3.**
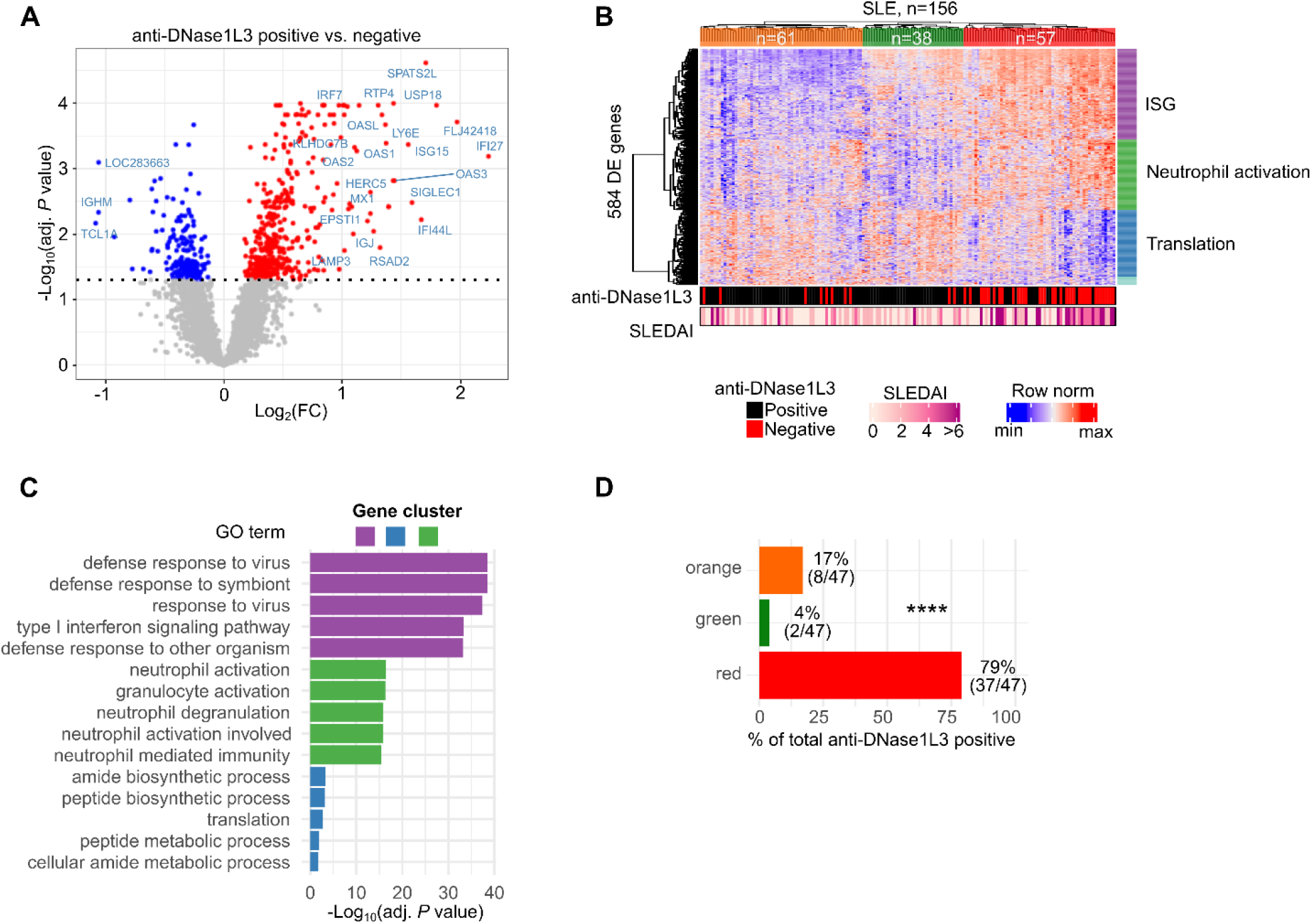
Transcriptional correlates of anti-DNase1L3 antibodies in SLE. (**A**) Volcano plot of 584 differentially expressed transcripts (DETs) between SLE patients positive and negative for anti-DNase1L3 antibodies. Red, 399 upregulated DETs with adjusted *P* < 0.01. Blue, 185 downregulated DETs with adjusted *P* < 0.01. (**B**). Hierarchical clustering of 584 DETs from **A**. Each column represents an individual patient and each row an individual gene. Top annotations show cluster membership. Bottom annotations indicate anti-DNase1L3 antibodies (positive = red, negative = black) and SLEDAI score in continuous scale. DETs were split in k=4 expression clusters and annotated by functional enrichment analysis using the g: Profiler toolset with the gene oncology molecular function (GO:MF) gene set collection. Red represents upregulated genes and blue downregulated genes. (**C**). Top 5 enriched GO:MF terms on gene expression clusters according to *P* value. (**D**) Frequency of anti-DNase1L3 antibody positive SLE patients according to cluster membership in **B**. GO, gene ontology; FC, fold-change. **** *P*< 0.0001.

**Figure 4.**
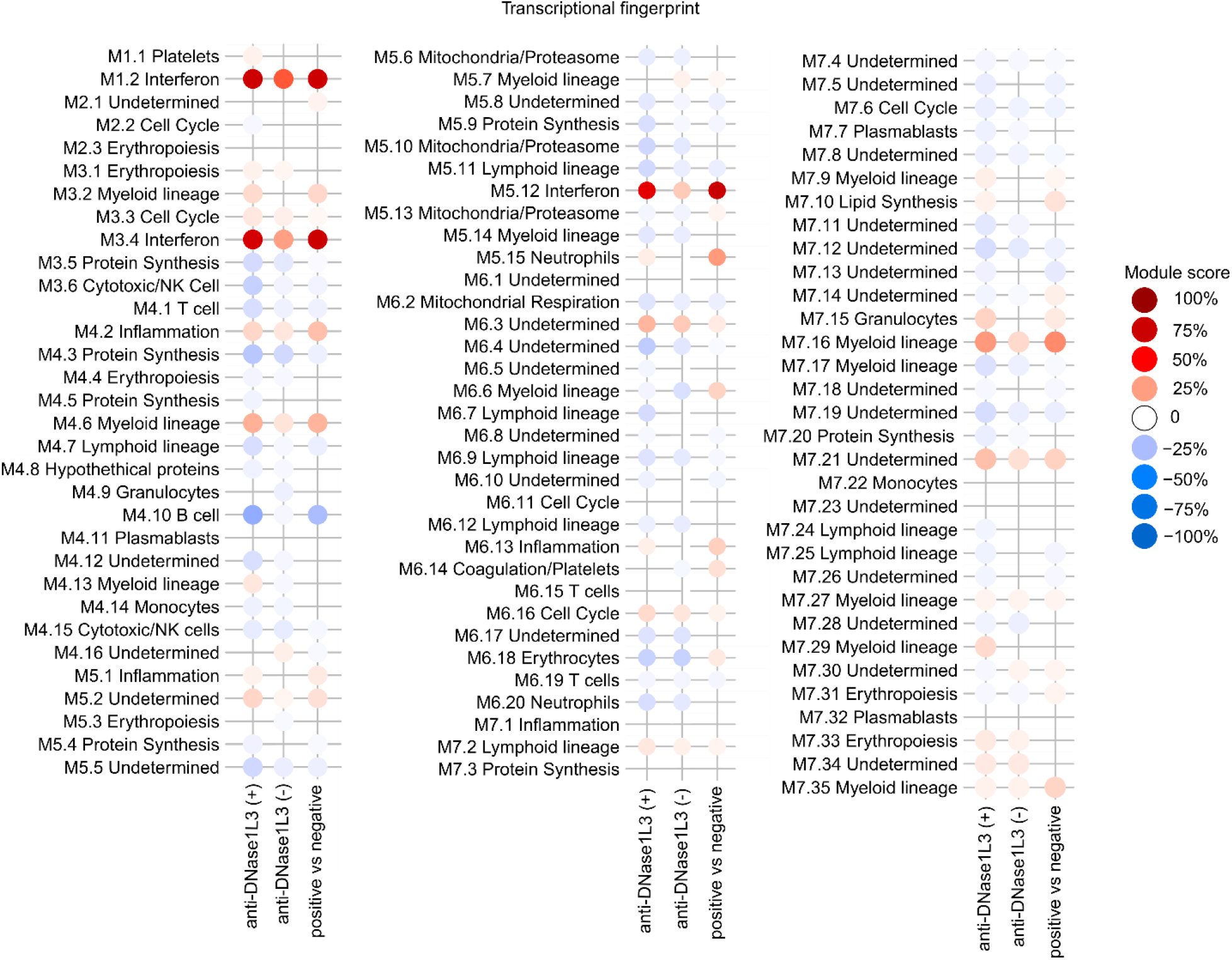
Transcriptional fingerprints associated with anti-DNase1L3 antibodies in SLE. Blood module fingerprints of SLE patients positive (+) and negative (-) for anti-DNase1L3 antibodies. Each dot represents a module, either overexpressed (red) or underexpressed (blue) as compared to healthy controls (left and middle columns), and anti-DNase1L3 positive *vs*. anti-DNase1L3 negative (right column). The intensity of the color represents the module score, which was calculated by the sum of upregulated (+1) and down regulated genes (−1) divided by the total number of transcripts within the module and expressed as a percentage. Only the modules with ≥5% or ≤ 5% of activity from the first 97 out of 260 modules are shown for clarity.

### Antibodies to DNase1L3 are associated with transcriptional fingerprints linked to disease activity in SLE

Patients with SLE display unique blood transcriptional profiles, including hallmark signatures linked to immune dysregulation (20). To define whether antibodies to DNase1L3 are associated with distinct transcriptional fingerprints in SLE, we used gene expression data from blood collected in parallel with the samples used to measure anti-DNase1L3 antibodies. We identified 584 differentially expressed transcripts (DETs) between anti-DNase1L3 negative and positive SLE patients (**Figure 3A**). Using unsupervised hierarchical clustering of the 584 DETs, SLE patients clustered in three major groups defined by the expression of IFN-stimulated genes (ISGs), inflammation, neutrophil activation genes, and genes related to mRNA processing and translation (**Figure 3, B and C**). Interestingly, 79% of patients positive for anti-DNase1L3 antibodies clustered with overexpression of ISGs and neutrophil activation genes, *P* <0.0001 (**Figure 3D**).

To further define whether anti-DNase1L3 antibodies associate with functional gene sets mechanistically linked to SLE (20), we conducted a modular analysis as described by Chaussabel et al (21). Interestingly, modules known to be overexpressed (i.e. the neutrophil, inflammation, cell cycle, and erythropoiesis modules) and underexpressed (i.e. NK cell/cytotoxicity, lymphoid lineage, B cells, T cells, and protein synthesis) in SLE (20) were associated with anti-DNase1L3 positivity (**Figure 4**). In particular, the IFN (i.e. M1.2, M3.4 and M5.12), myeloid/neutrophil/granulocyte (i.e. M3.2, M4.6, M5.15, M7.9, M7.15, M7.16 and M7.35) and inflammation (i.e. M4.2, M5.1 and M6.13) modules were the highest upregulated in the anti-DNase1L3 antibody positive subgroup (**Figure 4**). Interestingly, the IFN modules M1.2, M3.4 and M5.12 have been previously shown to correlate with SLE disease activity (22). Taken together, these data are consistent with clinical and laboratory features demonstrating that anti-DNase1L3 antibody positivity is associated with enhanced stimulation of immune pathways activated by cell-free DNA (23, 24).

### Neutralizing antibodies to DNase1L3 arise from autoreactive V_H_4-34-expressing IgG switched memory B cells

The autoantibody compartment in SLE is importantly shaped by the expansion of autoreactive B cells using the immunoglobulin variable heavy-chain gene segment V_H_4-34 (i.e. V_H_4-34^+^ cells) (25-29). To assess whether serum reactivity to DNase1L3 is linked to antibodies containing the V_H_4-34 encoded heavy chain, antibodies in serum were depleted using the anti-idiotypic antibody 9G4, a monoclonal antibody that specifically targets V_H_4-34 encoded antibodies (30). Interestingly, depletion of antibodies bearing the 9G4 idiotype decreased the reactivity to DNase1L3 by 36% to 80% in 5/6 anti-DNase1L3 positive SLE sera (**Figure 5A**), strongly supporting that a significant number of anti-DNase1L3 antibodies in SLE derive from autoreactive V_H_4-34-expressing B cells.

**Figure 5.**
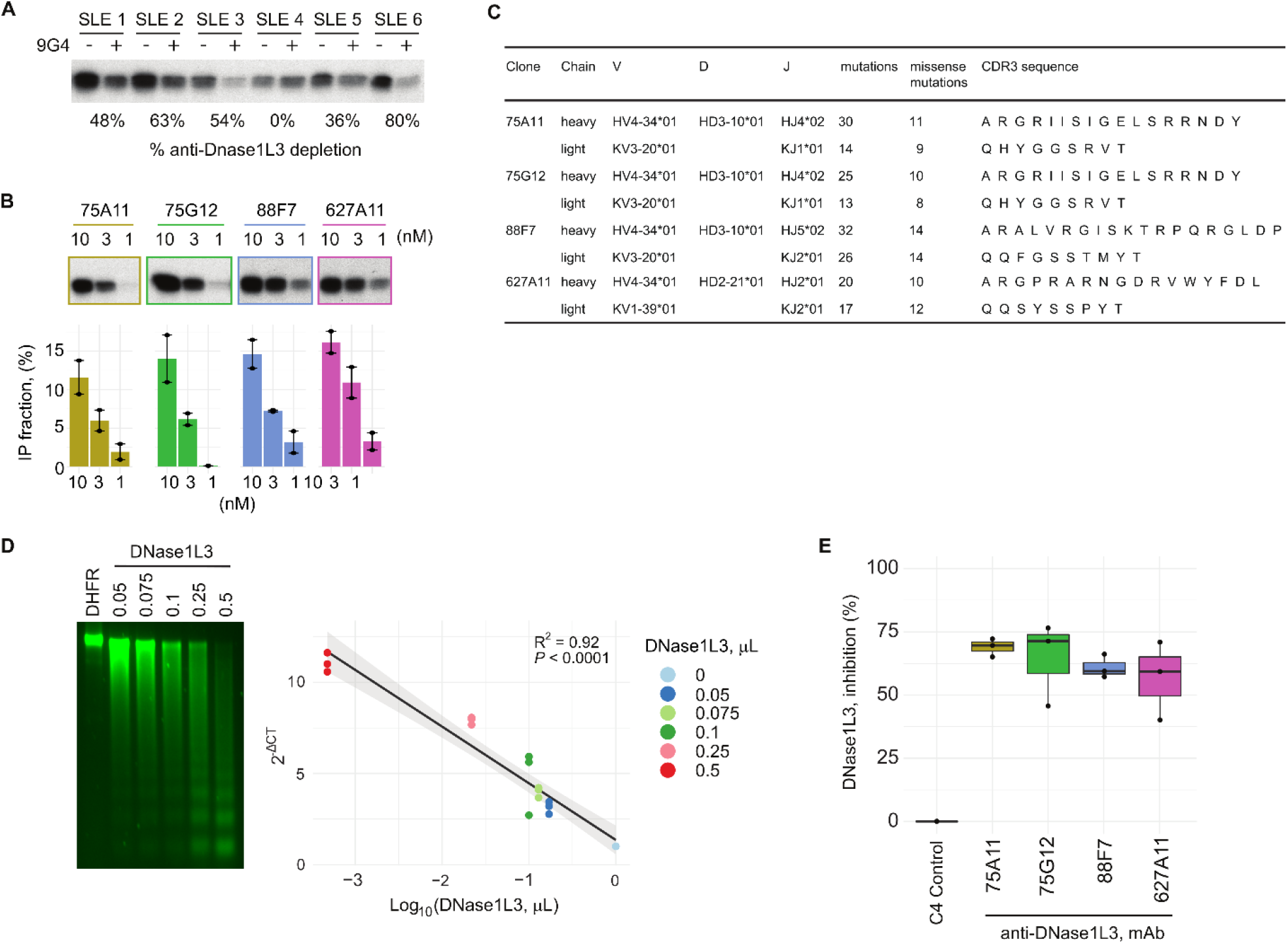
Origin and functional characterization of SLE patient-derived monoclonal antibodies to DNase1L3. (**A**) Radiolabeled DNase1L3 was immunoprecipitated (IP) with antiDNase1L3 positive SLE sera with (+) and without (-) immunoglobulin (Ig) depletion using 9G4 monoclonal antibodies. Radiolabeled DNase1L3 in immune complexes was quantified by densitometry and anti-DNase1L3 antibody depletion was expressed as percentage according to their corresponding 9G4 non-depleted serum. (**B**) Radiolabeled DNase1L3 was IP with increasing amounts of anti-DNase1L3 monoclonal antibodies (1-10 nM) (upper panel). The amount of DNase1L3 in immune complexes from **B** was quantified by densitometry and expressed as a fraction of the radiolabeled DNase1L3 input (100%). Mean (bar height), minimum and maximum (error bar) values are shown (lower panel). (**C**) Ig gene usage, mutation number and CDR3 amino acid sequences of monoclonal antibodies to DNase1L3. (**D**) Increasing amounts of recombinant DNase1L3 (PURExpress) were incubated with purified intact nuclei. Chromatin degradation was visualized in a 1.5% agarose gel (left panel) and quantified by LM-qPCR (right panel). Recombinant dihydrofolate reductase (DHFR, PURExpress) (1 µL) was used as negative control. (**E**) Effect of monoclonal antibodies (C4 control and anti-DNase1L3) on DNase1L3 activity. Chromatin degradation was quantified by LM-qPCR. The percentage of DNase1L3 inhibition was calculated as *DNase*1*L*3 _% *inh*_ = 1-2^Δ*ct*^ ⨯ 100, using the 2^-ΔCT^ from the conditions with DNase1L3 mAb with the C4 control as reference. Experiments were performed in two (**B** and **E**) and three (**D**) separate occasions.

To gain further insights into the origin and pathogenicity of anti-DNase1L3 antibodies in SLE, we screened for anti-DNase1L3 antibodies in a set of monoclonal antibodies previously generated from single B cells and antibody-secreting cells (ASCs) from SLE patients experiencing flares (28, 31). In particular, we initially focused on the analysis of 87 monoclonal antibodies largely generated from 9G4^+^ switched memory (SwM) B cells and ASCs (28, 31) (**Supplemental Table 2** and **Supplemental Figure 5**). Among these monoclonals, we found four antibodies with reactivity to DNase1L3 (i.e. 75G12, 75A11, 88F7, and 627A11) (**Figure 5B**), which were all derived from SwM B cells (**Supplemental Table 2** and **Supplemental Figure 5**). All antibodies were encoded by the self-reactive V_H_4-34 heavy chain variable region gene (**Figure 5C**). Based on the IgH V-D-J usage, complementarity-determining region 3 (CDR3) sequence, CDR3 length and the presence of common mutations, monoclonals 75G12 and 75A11 (isolated from the same patient) were determined to be clonally related (**Figure 5C**). The enrichment of somatic hypermutations (SHMs) in all antibodies supports that they originated from antigen-experienced cells.

The function of SLE patient-derived anti-DNase1L3 monoclonal antibodies was further addressed using recombinant DNase1L3 in chromatin digestion assays. Since DNase1L3 catalyzes DNA hydrolysis to produce broken ends that are blunt and 5’-phosphorylated (32), DNase1L3 activity was determined by absolute quantitation of inter-nucleosomally fragmented genomic DNA using ligation-mediated qPCR (LM-qPCR) (33, 34). First, we validated this assay by using increasing amounts of enzyme to digest native chromatin in intact nuclei (**Figure 5D**). Since DNase1L3 was generated using a cell-free transcription/translation system, dihydrofolate reductase (DHFR) expressed under similar conditions was used as negative control (**Figure 5D**). Then, using an enzyme concentration within the linear range of DNase1L3 activity, chromatin degradation was determined in the presence of monoclonal antibodies to DNase1L3. As a negative control, we used a monoclonal antibody generated from a single plasmablast (named C4) isolated from a healthy donor (**Supplemental Figure 6**). While the control monoclonal antibody had no effect on chromatin degradation mediated by DNase1L3, the monoclonal antibodies to DNase1L3 decreased DNA hydrolysis by the endonuclease by 60 to 70% (**Figure 5E**).

## DISCUSSION

Defective clearance of cellular debris remains as the strongest paradigm in the pathogenesis of SLE (35). Here, we found that patients with SLE have antibodies to DNase1L3, to date, the most important circulating endonuclease responsible for chromatin digestion from apoptotic and necrotic cells (10, 12). The production of autoantibodies to DNase1L3 is mediated by V_H_4-34^+^ cells, which are intrinsically autoreactive B cells that escape tolerance in SLE (36). Together, these findings provide new evidence supporting that pathways involved in the clearance of dying cells are targeted in SLE, and further identify the self-reactive encoding heavy-chain V_H_4-34 gene as a source of autoantibodies to DNase1L3 in this disease.

While the etiology of SLE is still unknown, a major advance in the study of SLE was the discovery that apoptosis is a major source of autoantigens in this disease (37-41). Understanding mechanisms that dysregulate the clearance of dying cells in SLE has therefore become a high priority. Under normal conditions, billions of cells die every day to maintain tissue homeostasis (42), and this process is importantly increased during inflammation, infection and cancer. Since the abnormal accumulation of dying cells can drive tissue damage and autoimmunity, unique mechanisms are responsible for the efficient clearance of cell remnants (35, 42). One of these mechanisms involves DNA endonucleases. These enzymes, called DNases, include the DNase1 and DNase2 family members, Caspase-activated DNase (CAD) and Endonuclease G. The DNase1 family has four members (DNase1, DNase1L1, DNase1L2, and DNase1L3) and the DNase2 family has three members (DNase2a, DNase2b and L-DNaseII) (7).

Overall, the absence of these enzymes either in humans or in mice leads to a wide variety of diseases (7). Among these, deficiency of DNase1 and DNase1L3 has been linked to SLE (43). DNase1 is the most widely expressed member of the DNase1 family, was the first DNase that gained pathogenic interest in SLE (7, 43, 44), and was found to be a target of autoantibodies in a subset of patients with SLE (45). The interest on DNase1 in SLE, however, likely declined because of its failure to show clinical benefit in a phase Ib trial using recombinant human DNase1 in patients with lupus nephritis (46). Since DNase1L3 is more efficient than DNase1 in the inter-nucleosomal cleavage of genomic DNA, it is rational to suggest that DNase1L3 may have a more critical role in the degradation of extracellular chromatin and therefore, in regulating the load of immunogenic DNA. This notion is supported by the finding that DNase1L3 deficiency is associated with a prominent antibody response to dsDNA and the development of SLE, both in humans and mice (5, 11, 14). In the absence of DNase1L3 activity, lupus development is explained by increased availability of antigenic cell-free DNA, driving TLR-dependent IFN-I production and the induction of antibodies to dsDNA (16). Similar to DNase1L3 deficiency, our study demonstrates that the presence of anti-DNase1L3 antibodies in patients with SLE is strikingly associated with features suggesting an abnormal immune response to DNA. These include a prominent association with anti-dsDNA antibodies and with immune pathways activated by cell-free DNA protein complexes (e.g. containing HMGB1 or anti-dsDNA antibodies), such as neutrophil activation (23) and the overexpression of ISGs (24), known as the “interferon signature” (47).

Interestingly, during the final preparation of this manuscript, another group reported the presence of anti-DNase1L3 antibodies in SLE, which associated with lupus nephritis and the accumulation of cell-free DNA in circulating microparticles (18). Our work validates the existence of anti-DNase1L3 antibodies in an independent cohort, while significantly extending our understanding on the immunological basis of the origin and pathogenic effects of these autoantibodies and providing new insights into potential therapeutic interventions. Although our work and the study published by Hartl et al (18) identified the presence of antibodies to DNase1L3 in SLE, these studies have some significant differences that are important to address. Quantifying the circulating activity of DNase1L3 based on chromatin degradation assays encounters numerous challenges. In the study by Hartl et al (18), DNase1L3 activity was determined in plasma, but there was no specification of the type of anticoagulant that was used (heparin, EDTA or citrate) and how the caveats of using plasma were overcome, in particular, those that may facilitate chromatin degradation by DNase1 (i.e. heparin-induced displacement of histones from chromatin, DNase1L3 inhibition, and activation of the coagulation pathway as result of plasma recalcification) (10). To circumvent these problems, we avoided using plasma and instead, serum was treated with aprotinin to inhibit serine proteases.

While these measures can increase the specificity of the assay for DNase1L3 activity, it is important to recognize that serum/plasma from SLE have additional components that can affect chromatin degradation, which are difficult to control. First, several autoantibodies in SLE target chromatin components (e.g. anti-dsDNA, anti-chromatin and/or anti-histones), which can affect substrate accessibility by the enzyme, blocking chromatin breakdown by endonucleases (48). Thus, the presence of different levels and types of autoantibodies to elements in chromatin are likely to affect the results of this assay. Second, increased production of DNase1L3 by activated immune cells (9) may overcome circulating inhibition of the enzyme by neutralizing antibodies to DNase1L3. Because of the numerous technical difficulties, we consider that chromatin degradation is neither a feasible assay to precisely quantify DNase1L3 activity, nor a good biomarker to correlate with the presence of anti-DNase1L3 antibodies in SLE. Instead, our data demonstrate that the direct detection of anti-DNase1L3 antibodies seems a better strategy to identify patients in which the DNase1L3 pathway is pathogenically targeted in sporadic SLE. In particular, our data shows that positivity for anti-DNase1L3 antibodies distinguishes a subset of SLE patients characterized by systemic IFN and myeloid/neutrophil activation, anti-dsDNA antibodies, complement activation, and higher disease activity.

The functional analysis of anti-DNase1L3 antibodies in SLE is also challenging. In the study by Hartl et al (18), bulk IgG purified from SLE plasma was used to study the effect of these antibodies on DNase1L3 activity. However, purified IgG is highly enriched with numerous other autoantibody specificities that may affect chromatin degradation assays. Thus, it is difficult to attribute the effect of bulk IgG on DNase1L3 activity to a single antibody specificity (i.e. anti-DNase1L3 antibodies) considering the extensive pool of autoantibodies found in SLE (49). Therefore, we focused on the identification of SLE patient-derived monoclonal antibodies to DNase1L3, which can directly address the specific function of these antibodies and additionally provide insights into their origin and genetic characteristics. Coincidently, Hartl et al (18) also studied some SLE-derived 9G4^+^ monoclonal antibodies obtained from the same source (28, 31), but these antibodies were used to address binding to microparticles and not the effect on DNase1L3 specificity.

Within a large panel of monoclonal antibodies generated from SLE patients experiencing flares, we identified four anti-DNase1L3 antibodies (two were clonally related) derived from autoreactive V_H_4-34^+^ SwM B cells, which all decrease chromatin degradation by DNase1L3. B cells bearing the 9G4 idiotype (encoded by the V_H_4-34 heavy chain) are inherently autoreactive cells regulated by peripheral checkpoints (36, 50). In healthy individuals, these cells are largely excluded from the germinal centers and underrepresented in the memory compartment (50). In patients with SLE, however, 9G4-B cells progressed through this checkpoint and successfully participate in germinal center reactions, generating increased levels of IgG memory and plasma cells (36). Indeed, 9G4 antibodies represent 10–45% of the total serum IgG in patients with active disease (25-27, 36).

In summary, this study demonstrates that anti-DNase1L3 antibodies represent a new subset of autoantibodies linked to the expansion of autoreactive VH4-34^+^ B cells in SLE. Moreover, the data provide evidence for a mechanistic association between anti-DNase1L3 antibodies and immune pathways activated by immunogenic self-DNA, such as the IFN and myeloid/neutrophil signatures, and the production of antibodies to dsDNA. Importantly, the finding that autoreactive B cells bearing the 9G4 idiotype encode for multiple autoantibody specificities (28, 29), including anti-DNase1L3 antibodies, highlights VH4-34^+^ B cells and 9G4-antibodies as potential targets for the treatment of SLE.

## MATERIALS AND METHODS

### Participants

Stored PBMC RNA from 34 consecutive SLE patients and 12 healthy controls were used to examine DNase1L3 expression by qPCR. In addition, DNase1L3 activity was quantified in 20 SLE patients and 10 healthy controls in which sera collected in parallel with RNA PBMCs were also available. Sera from a convenience cohort of patients with active SLE (n = 24) and healthy controls (n = 22) were used as anti-DNase1L3 antibody discovery cohort. Sera from 62 healthy controls and 158 SLE patients from the “Study of biological Pathways, Disease Activity and Response markers in patients with Systemic Lupus Erythematosus” (SPARE) (51) cohort were studied to define the prevalence and clinical significance of antibodies to DNase1L3 in SLE. SPARE is a prospective observational cohort that has been extensively described previously (19, 51). Briefly, adult patients (age 18 to 75 years-old) who met the definition of SLE per the revised American College of Rheumatology classification criteria were eligible into the study (52). At baseline, the patient’s medical history was reviewed, and information on current medications was recorded. Patients were followed-up over a 2-year period. Patients were treated according to standard clinical practice. Disease activity was assessed using the Safety of Estrogens in Lupus Erythematosus: National Assessment (SELENA) version of the Systemic Lupus Erythematosus Disease Activity Index (SLEDAI) (53) and physician global assessment (PGA) (54). C3, C4, anti-dsDNA (Crithidia), complete blood cell count and urinalysis were performed at every visit. Study participants also underwent whole blood gene expression analysis at baseline using the Affymetrix GeneChip HT HG-U133+ (19, 51). Eighty-seven monoclonal antibodies previously generated from SLE patients experiencing flares were used to screen for anti-DNase1L3 antibodies (28, 31). All samples were obtained under approval by the Institutional Review Boards at the Johns Hopkins University School of Medicine and Emory University School of Medicine.

### DNase1L3 activity in serum

Serum activity of DNase1L3 was determined as previously described (10) with some modifications. Briefly, serum at 50 µL/mL in DNase reaction buffer (10 mM Tris/HCl, 50 mM NaCl, 2 mM MgCl_2_, 2 mM CaCl_2_, pH 7.0) was incubated with recombinant aprotinin (Sigma) at 500 U/mL. After 10 mins, nuclei purified from Expi293 cells (isolated as described) (25) were added at 5×10^5^/mL and further incubated for additional 2 hrs at 37°C. The final volume of each reaction was 20 µL. Reactions were stopped by adding 20 µL of proteinase K buffer (PKB, 50 mM Tris-HCl pH 8.0, 1 mM EDTA, 2.5% Tween 20, and 800 Units/mL proteinase K) and incubation at 50°C for 20 min, followed by heat inactivation 95°C for 5 min (55). After centrifugation at 10,000xg, the amount of remaining DNA was measured by qPCR with primers for human Alu repeats (55). qPCR amplification was performed using iTaq Universal SYBR Green Supermix (Bio Rad) as follow: 95°C for 4 min (1 cycle), followed by 30 cycles of denaturation at 95°C for 15 s, annealing at 64°C for 30 s, and extension at 72°C for 30 s (55). The remaining DNA was estimated using a calibration curve of serial dilutions of DNA. The fraction of degraded chromatin was calculated by subtracting the remaining DNA from the total DNA input and expressed as arbitrary units, where 0 equals to no degradation and 1 to full degradation of chromatin.

### DNase1L3 mRNA expression

Total RNA was purified from PBMCs using the RNeasy Plus Mini Kit (Qiagen). cDNA was synthesized using SuperScript IV VILO Master Mix (Invitrogen). qPCR was performed using TaqMan primers and probes (Applied Biosystems) to assess the expression of DNase1L3 (Hs00172840_m1) and 18S was used as normalizer gene. The relative expression was calculated with the 2^−ΔΔC^_T_ method (56).

### DNase1L3 cloning and protein expression

cDNA encoding human mature DNase1L3 (amino acids 21-305) was synthesized using RNA from human PBMCs and cloned into pcDNA3.1 and pET-28a(+). In pET-28a, the 5’ site end in mature DNase1L3 was cloned at the NcoI site in the vector. Thus, the T7 promoter is followed by a ribosome binding site and the DNase1L3 start codon. The protein encoded by pET-28a-DNase1L3 is not tagged. pcDNA3.1-DNase1L3 was used to generate ^[35S]^methionine-labeled DNase1L3 by TNT T7 Quick Coupled Transcription/Translation (Promega). pET-28a-DNase1L3 was used to generate recombinant active DNase1L3 using the PURExpress In Vitro Protein Synthesis Kit (New England Biolabs). The control plasmid encoding dihydrofolate reductase (DHFR) is included in the PURExpress kit.

### Immunoprecipitation

Radiolabeled DNase1L3 was immunoprecipitated with 2 μl of serum, 10 µL of cell supernatants from anti-DNase1L3 antibody expressing cells, or with purified anti-DNase1L3 monoclonal antibodies in 300 µL of NP-40 buffer (20 mM Tris/HCl, 150 mM NaCl, 1 mM EDTA, 1% Nonidet P40, pH 7.4) for 1 hr at 4°C. Protein A beads were added and incubated for additional 30 min at 4°C. After three washes with vortexing in NP-40 lysis buffer, beads were boiled in SDS sample buffer. Samples were separated by gel electrophoresis, and immunoprecipitated proteins were visualized by radiography. Densitometry was performed on all films and values were normalized to a high-titer anti-DNaseL13 serum. Antibody positivity was defined using a cutoff of two standard deviations above the mean anti-DNase1L3 antibody level in healthy sera.

### Production and purification of monoclonal antibodies

The cloning of monoclonal antibodies from single B cells and antibody-secreting cells (ASCs) from SLE patients experiencing flares was previously described (28, 31). Single peripheral blood plasmablasts (CD3-CD14-CD19+CD20-CD27+CD38+) from a healthy female donor were sorted into 96-well PCR plates. IgH and IgL (κ or λ) variable regions were cloned into expression vectors containing human Igγ1, Igκ, or Igλ constant regions as previously described (57). One monoclonal antibody (C4) was highly expressed and was selected for further characterization. Analysis of the antigen specificity of C4 was performed using the HuProt human proteome microarray (CDI). C4 exhibited broad, non-specific reactivity to 0.25% of the >21,000 proteins on the array, but had no reactivity to DNAse1L3. This antibody was used a monoclonal control. 293T cells in high glucose DMEM and 10% ultra-low IgG fetal bovine serum (Gibco) were co-transfected with plasmids encoding IgH and IgL using Expifectamine 293 transfection reagent (Gibco). ExpiFectamine 293 transfection enhancers (Gibco) were added 20 hrs after plasmid transfection. Supernatants were collected at day 5 after transfection and the antibodies purified using Protein A beads (Pierce).

### Quantitation of inter-nucleosomally fragmented genomic DNA using ligation-mediated qPCR (LM-qPCR)

DNA degradation was measured by quantitation of inter-nucleosomally fragmented DNA by LM-qPCR as previously described (34) with some modifications. Briefly, the DHApo1 and DHApo2 oligonucleotides (5’ to 3’: AGCACTCTCGAGCCTCTCACCGCA and TGCGGTGAGAGG, respectively) were annealed by mixing 50 µL (100 pmol/μl) of each in 250 µl of 250 mM Tris (pH 7.7), heating the mixture to 90°C for 5 min, incubating at 55°C for 15 min, and allowing the mixture to cool to RT. The linker mixture was frozen and thawed on ice before use. Ligation reactions (20 µL) were performed overnight at 16°C using Quick T4 ligase (New England Biolabs), 100 ng DNA and 1 µL linker. qPCR was performed using the DHApo1 primer and iTaq Universal SYBR Green Supermix (Bio Rad) as follow: 95°C for 4 mins (1 cycle), 72°C for 4 mins (1 cycle), followed by 40 cycles of denaturation at 94°C for 1 min and annealing/extension at 72°C for 3 mins.

### Neutralizing activity of monoclonal antibodies to DNase1L3

DNase1L3 and control DHFR were generated using the PURExpress In Vitro Protein Synthesis Kit (NEB). DNase1L3 activity was titrated by co-incubating increasing amounts of PURExpress DNase1L3 (0.05 µL, 0.075, 0.1 µL, 0.25 and 0.5 µL) with 10,000 purified nuclei in 20 µL of DNase reaction buffer containing 5% bovine serum albumin (BSA) solution (Sigma). PURExpress DHFR was used as negative control. After 30 mins at 37°C, reactions were stopped by adding 20 µL of PKB, incubation at 50°C for 20 mins and heat inactivation at 95°C for 5 min. DNA was purified by isopropanol precipitation and resolved in a 1.5% agarose gel. In addition, DNA degradation was measured by quantitation of inter-nucleosomally fragmented DNA by LM-qPCR. The amount of inter-nucleosomally fragmented DNA was calculated by the 2^-Δ*ct*.^method, using the condition with PURExpress DHFR as reference. No DNase activity was found in the PURExpress synthesis kit unless DNase1L3 was expressed (**Figure 5C**). To assess the effect of anti-DNase1L3 monoclonal antibodies on DNase1L3 activity, antibodies at 1.6 µM were incubated with 0.1 µL of PURExpress DNase1L3 in DNase reaction buffer containing 5% BSA. The monoclonal antibody C4 was used as antibody control. In addition, purified nuclei were incubated with PURExpress DHFR as control for undigested DNA. After 1 hr at room temperature (RT), 10,000 purified nuclei were added and further incubated for 30 mins at 37°C. The final volume of the reaction was 20 µL. Reactions were stopped with PKB as described above and DNA was purified by isopropanol precipitation. Inter-nucleosomally fragmented DNA was quantified by LM-qPCR. The inter-nucleosomally fragmented DNA was calculated by the 2^-Δ*ct*.^ method with C4 control as reference. The percentage of DNase1L3 inhibition was calculated as *DNase*1*L*3_%*inh*_ = 1-2^-Δ*ct*^ ⨯ 100

### Serum depletion of antibodies bearing the 9G4 idiotype

Sera depletion was performed using the Pierce™ Protein G IgG Plus Orientation Kit (Thermo) primarily to manufacture suggestions. The idiotypic rat anti-human 9G4 mAb was saturated within the agarose and cross-linked utilizing a multi-flow through load. 500µL of patient sera was utilized and bound overnight at 4°C prior to flow through. The elution protocol and column wash resulted in roughly a 1:5 dilution of the sera volume following flow through. Total IgG and 9G4 specific ELISAs were performed on the sera pre- and post-depletion. The total IgG loss ranged from 10-27% following the 9G4 column depletion, however the 9G4 IgG loss was greater than 99% in all samples.

### Gene expression analyses

Gene expression analysis from the SPARE cohort was previously described (19). CEL files were subjected to RMA background correction, and quantile normalization, using the Oligo package (20). To select only expressed genes in whole blood, we filtered out transcripts that had a raw signal < 100 in less than 10% of samples with the genefilter R package. All calculations and analyses were performed using R (ver 4.0.2) and Bioconductor (ver 3.13) (58). Differentially expressed transcripts (DETs) were analyzed using the R package limma (59). Functional gene set enrichment was carried out with the R interface gprofiler2 for the server g:Profiler (60).

### Calculation of the blood expression modules activity

DETs were analyzed using the R package limma (59). The Blood gene expression modules from Chaussabel et. al.(21) were obtained from the R package “tmod” for Bioconductor (61). To calculate module activity at the group level, DETs were obtained by comparing SLE anti-DNase1L3 positive and negative subgroups against healthy controls, and by comparing SLE anti-DNase1L3 positive *versus* antiDNse1L3 negative. Then, each DET was coded as 1 when upregulated, −1 when downregulated, and 0 when not significantly regulated. Module activity was calculated by addition of the coded value of all DET within a module divided by the total number of genes in the module. Module activity was expressed as a percentage.

### Statistical analyses

Comparisons of continuous variables between groups were done using Student’s T test and ANOVA test as indicated. The Mann-Whitney’s U test and KruskalWallis test was used for group-wise comparisons of non-normally distributed variables Fisher’s exact test and χ^2^ tests were used for univariate analysis on SPARE cohort variables, as appropriate. Exact2×2 package in R version 3.5.1 was used for binary variables to obtain p-value, OR, and 95% CI. Unsupervised hierarchical clustering with complete linkage was performed by computing a correlation-based distance between genes (Pearson’s method) and the Canberra metric for the distance between subjects. Heatmap visualization was done using the Complex heatmap R package (62). To improve visualization, dendrograms were reordered using the modular leaf ordering methods from the dendsort R package (63). Statistical significance was set at p < 0.05. The statistical analyses were carried with the R software version 4.0.2 and SPSS IBM statistics version. 25.

## Data Availability

Data or materials derived from human samples may be requested subject to any underlying restrictions on such data or samples, and will require material transfer and data use agreements through the Johns Hopkins University School of Medicine or Emory University School of Medicine.

## Author contributions

Conception: FA. Designing research studies: YY, EG-B, JL, KSC, MIT-Z, RB, YW, ASC, MP, IS, FA. Conducting experiments: YY, EG-B, KSC, RB, YW, ASC, MPz, DPF, MIT-Z, FA. Data acquisition and analyses: YY, EG-B, JL, KSC, RB, YW, ASC, MPz, MIT-Z, DPF, DWG, ED, MP, IS, FA. Interpretation of data: YY, EG-B, JL, KSC, ED, MP, IS, FA. Providing reagents: CAS-B, MP, ED, IS. Writing—original draft: EG-B, F.A. Writing—review and editing: YY, EG-B, JL, KSC, RB, YW, ASC, MP, MIT-Z, DPF, CAS-B, DWG, ED, MP, IS, FA. All authors reviewed, edited, and approved the manuscript.

## Acknowledgments

We would like to thank the support of all members of the Petri, Sanz, and Andrade labs. Special thanks to Dr. S. Sam Lim and the Grady Memorial Hospital in Atlanta, as well as Emory University Hospital, and the University of Rochester Medical Center for sample support, and all the willing donors from the SPARE Cohort and from all contributing institutions. This project was supported by the Rheumatology Research Foundation, the Jerome L. Greene Foundation, the National Institute of Arthritis and Musculoskeletal and Skin Diseases (NIAMS), and the National Institute of Allergy and Infectious Diseases (NIAID) at the National Institutes of Health (NIH) grants number R01 AR069569, R21 AI147598, R01 AR069572, P01 AI125180, R37 AI049660, U19 AI110483, and the Intramural Research Program of the National Institute on Aging to Ranjan Sen. The contents of this article are solely the responsibility of the authors and do not necessarily represent the official views of the NIH, NIAMS or NIAID. YY received a scholarship from The Wuhan Scientific Funding for young investigators: 201271031432.

