## Supplemental material for "Neutralizing anti-DNase1L3 antibodies derive from autoreactive VH4-34^+^-B cells and associate with the interferon signature in SLE"

**Contents**

**Supplemental Figure 1.** Detection of anti-DNase1L3 antibodies in sera from the SPARE cohort and healthy controls.

**Supplemental Figure 2.** Current treatment of SLE patients according to anti-DNase1L3 status.

**Supplemental Figure 3.** Trajectories of anti-DNase1L3 levels and disease activity in SLE anti-DNase1L3 positive patients.

**Supplemental Figure 4.** Trajectories of anti-DNase1L3 levels and disease activity in SLE anti-DNase1L3 negative patients.

**Supplemental Figure 5.** Identification of SLE patient-derived monoclonal antibodies to DNase1L3.

**Supplemental Figure 6.** Ig gene usage, mutation number and CDR3 amino acid sequences of monoclonal antibody C4.

**Supplemental Table 1.** Demographics and clinical characteristics of SLE patients according to anti-DNase1L3 antibody.

**Supplemental Table 2.** SLE patient-derived monoclonal antibodies.

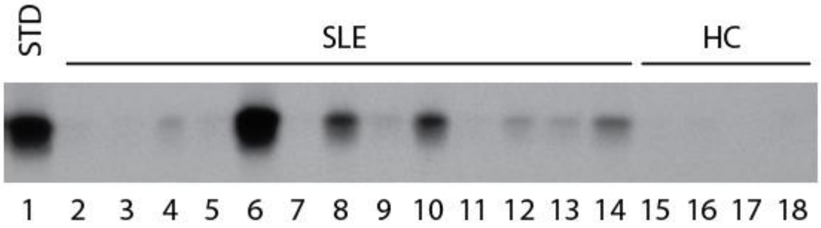


**Supplemental Figure 1. Detection of anti-DNase1L3 antibodies in sera from the SPARE cohort and healthy controls.** Sera from the SPARE cohort and healthy controls (HC) were used to immunoprecipitate radiolabeled DNase1L3. Anti-DNase1L3 units were calculated by densitometry and the values were normalized to a high-titer anti-DNaseL13 serum (standard, STD). Shown is a representative radiograph including the STD, 13 SPARE, and 4 HC sera.

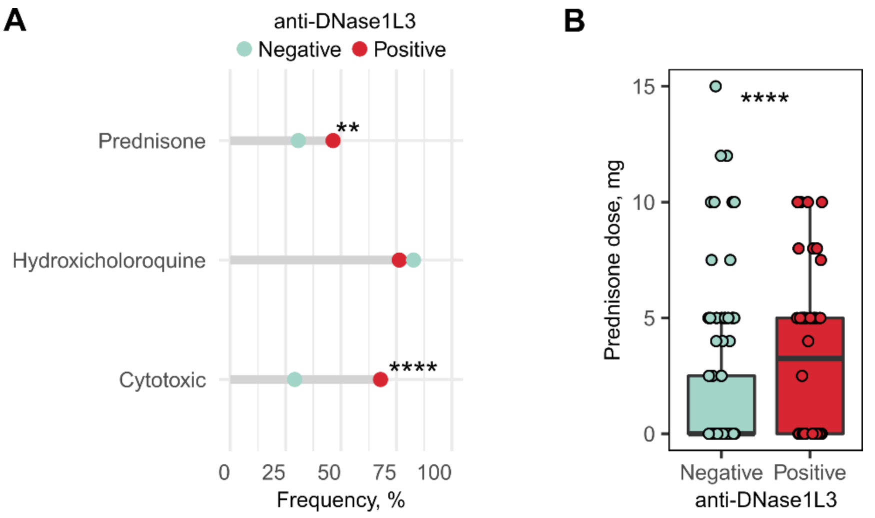


**Supplemental Figure 2. Current treatment of SLE patients according to anti-DNase1L3 status.** **(**A) Current treatment at the time of visit of SLE subjects according to anti-DNase1L3 status. **(B)** Prednisone dose (mg) at the visit of SLE subjects according anti-DNase1L3 status. Comparisons between groups were done using Fisher's exact test or Mann-Whitney's U, accordingly. ****$P$ < 0.0001, ***$P$ < 0.001 and **$P$ < 0.01.

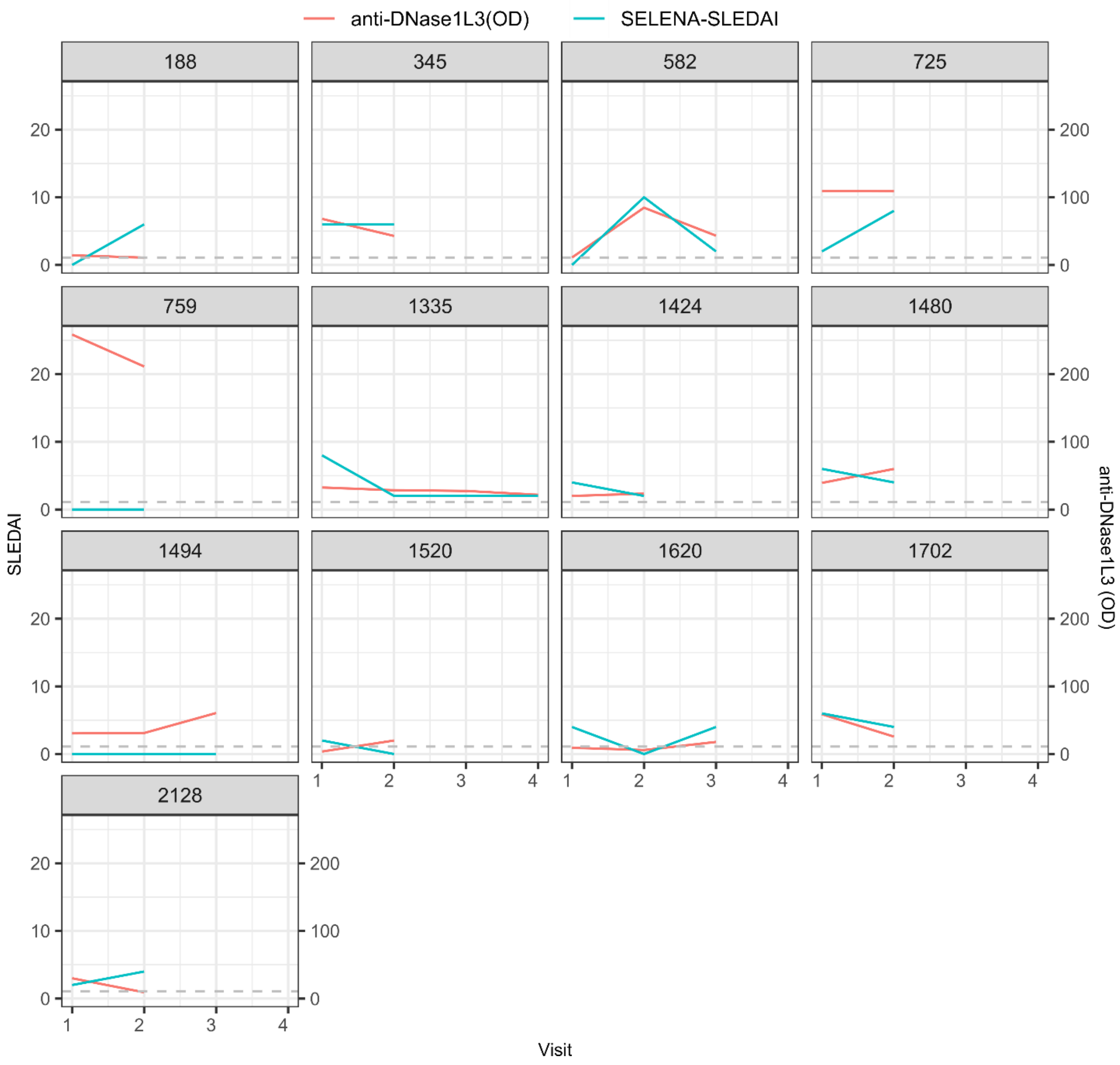


**Supplemental Figure 3. Trajectories of anti-DNase1L3 levels and disease activity in SLE anti-DNase1L3 positive patients.** Disease activity scores (SLEDAI) and anti-DNase1L3 antibody levels were longitudinally evaluated in 13 patients with SLE who were positive for anti-DNase1L3 antibodies in at least one visit.

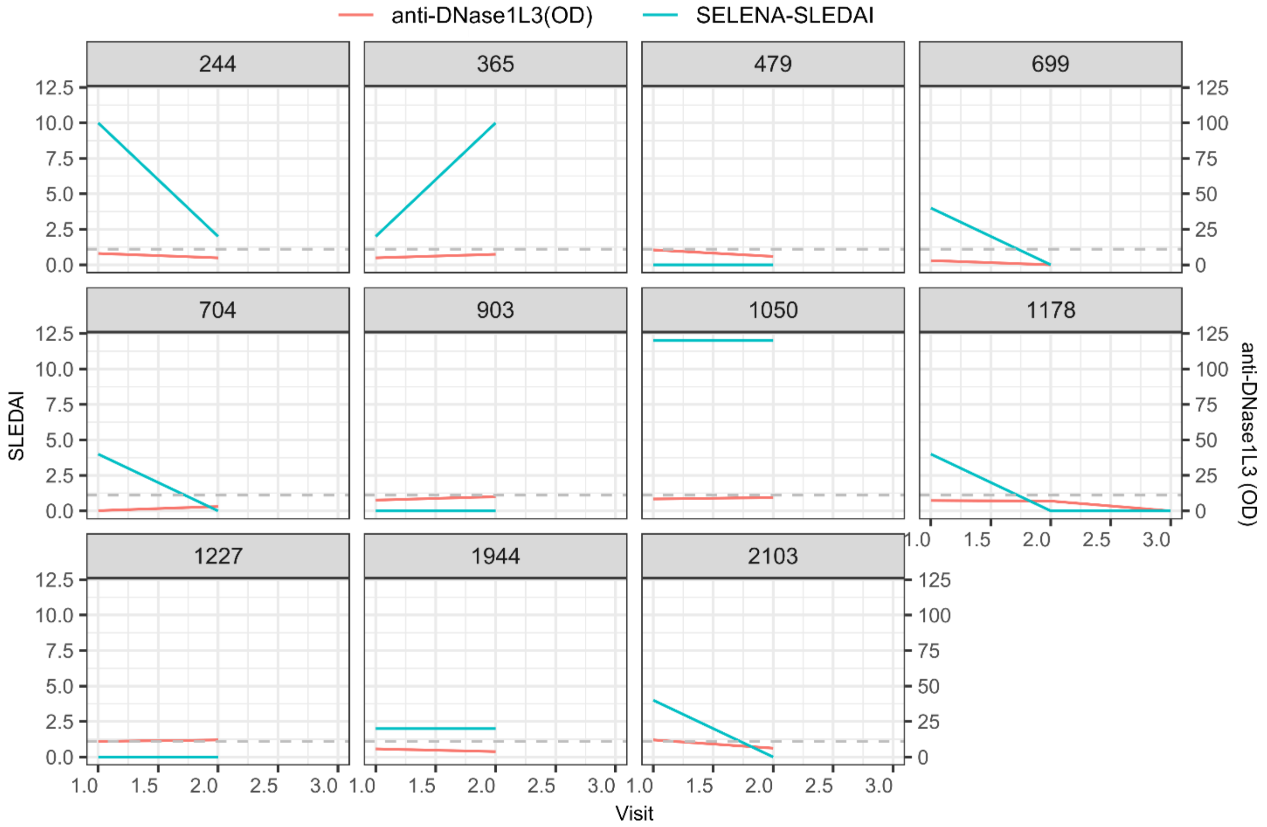


**Supplemental Figure 4. Trajectories of anti-DNase1L3 levels and disease activity in SLE anti-DNase1L3 negative patients.** Disease activity scores (SLEDAI) and anti-DNase1L3 antibody levels were longitudinally evaluated in 11 patients with SLE who were negative for anti-DNase1L3 antibodies in every visit.

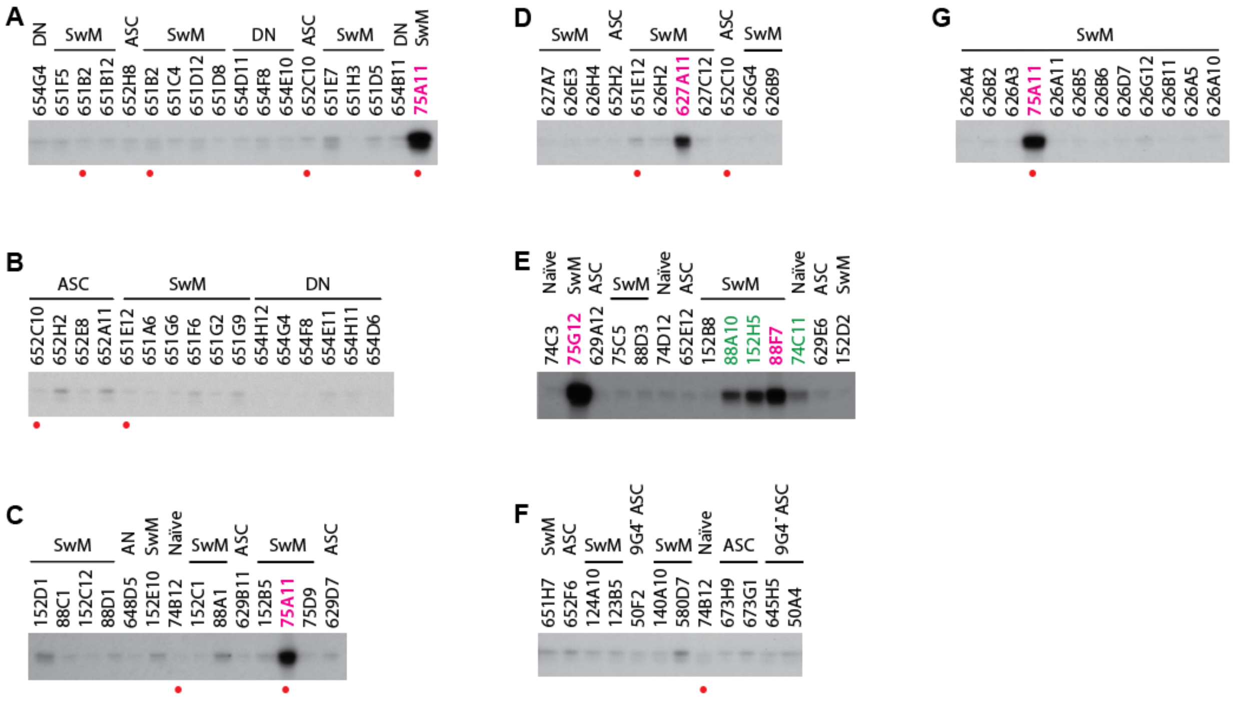


**Supplemental Figure 5. Identification of SLE patient-derived monoclonal antibodies to DNase1L3.** (**A-G**). Supernatants from 293 cells expressing monoclonal antibodies generated from single B cells and antibody-secreting cells (ASCs) from SLE patients experiencing flares were screened by immunoprecipitation using radiolabeled DNase1L3. Among seven antibodies with reactivity to DNase1L3, three antibodies (panel **E**, marked in green) were not verified using affinity purified antibodies. Antibodies with confirmed reactivity to DNase1L3 are marked in pink. Antibodies marked with a red dot were unintentionally screened more than once. Except for clones 50F2, 645H5 and 50A4 (panel **F**), the rest of monoclonal antibodies are from $9G4^{+}$ cells. SwM, switched memory. DN, double negative.

| Clone | Chain | V | D | J | Mutations | Missense mutations | CDR3 sequence |
|---|---|---|---|---|---|---|---|
| C4 | Heavy | HV3-49*03 | HD2-2*01 | HJ4*02 | 11 | 4 | TRNRPGYCSGTSCCLD |
| | Light | KV2-28*01 | | KJ4*01 | 7 | 4 | MQALQIPQT |

**Supplemental Figure 6.** Ig gene usage, mutation number and CDR3 amino acid sequences of monoclonal antibody C4.

**Supplemental Table 1**. Demographics and clinical characteristics of patients by Anti-DNase1L3 antibody

| | Anti-DNase1L3 Positive n = 48 (100%) | Anti-DNase1L3 Negative n = 110 (100%) | OR (95% CI) | *P* value |
|---|---|---|---|---|
| ***Demographic characteristics*** | | | | |
| Female | 47 (97.9%) | 102 (92.7%) | 3.7 (0.45,30.32) | 0.278 |
| Ethnicity | | | | 0.669 |
| Black | 18 (37.5%) | 45 (40.9%) | 0.9 (0.45,1.91) | |
| White | 25 (52.1%) | 58 (52.7%) | 1 | |
| Other | 5 (10.4%) | 7 (6.4%) | 1.7 (0.48,5.73) | |
| Ever Smoker, n (%) | 19 (39.6%) | 42 (38.2%) | 1.1 (0.53,2.12) | 0.861 |
| Alcohol abuse | 3 (6.3%) | 10 (9.1%) | 0.7 (0.18,2.54) | 0.756 |
| Drugs | 2 (4.2%) | 7 (6.4%) | 0.6 (1.13,3.20) | 0.724 |
| ***Clinical characteristics (Ever)*** | | | | |
| Livedo | 23 (47.9%) | 33 (30%) | 2.1 (1.06,4.35) | 0.046 |
| Proteinuria | 29 (60.4%) | 46 (41.8%) | 2.1 (1.03,4.37) | 0.038 |
| Mono multiplex | 2 (4.2%) | 0 (0%) | N/A | 0.091 |
| Anemia | 42 (87.5%) | 75 (68.2%) | 3.3 (1.27,8.51) | 0.011 |
| Leukopenia | 27 (56.2%) | 43 (39.1%) | 2.0 (0.97,4.03) | 0.056 |
| Lymphopenia | 26 (54.2%) | 42 (38.2%) | 1.9 (0.92,3.82) | 0.081 |
| Moon Facies | 23 (47.9%) | 34 (30.9%) | 2.1 (1.02,4.15) | 0.048 |
| Cataracts | 11 (22.9%) | 48 (43.6%) | 0.4 (0.17,0.83) | 0.020 |
| Other arterial thrombosis | 5 (10.4%) | 3 (2.8%) | 4.1 (0.95,20.31) | 0.058 |
| Herpes zoster | 17 (35.4%) | 23 (20.9%) | 2.1 (0.97,4.63) | 0.073 |
| Splenomegaly | 2 (4.2%) | 0 (0%) | N/A | 0.091 |
| Cytotoxic use ever | 38 (79.2%) | 61 (55.5%) | 3.0 (1.37,6.92) | 0.005 |
| Lupus anti-coagulant | 23 (47.9%) | 31 (28.2%) | 2.3 (1.15,4.81) | 0.019 |
| Anti-cardiolipin | 39 (81.2%) | 65 (59.1%) | 3.0 (1.3,7.05) | 0.010 |
| Anti-B2 Glycoprotein | 22 (45.8%) | 27 (25%) | 2.5 (1.21,5.35) | 0.015 |
| Anti-Sm | 11 (22.9%) | 21 (19.1%) | 1.3 (0.51,2.92) | 0.668 |
| FP-RPR | 7 (14.6%) | 11 (10%) | 1.5 (0.49,4.37) | 0.423 |
| ANA | 48 (100%) | 109 (99.1%) | N/A | 1.000 |
| Anti-dsDNA | 41 (85.4%) | 58 (52.7%) | 5.2 (2.09,13.54) | < 0.001 |
| Anti-Ro | 22 (45.8%) | 27 (24.5%) | 2.6 (1.24,5.48) | 0.009 |
| Anti-La | 9 (18.8%) | 15 (13.6%) | 1.5 (0.54,3.69) | 0.471 |
| Anti-RNP | 18 (37.5%) | 24 (21.8%) | 2.1 (0.99,4.71) | 0.051 |
| Low CH50 | 15 (31.2%) | 8 (7.3%) | 5.7 (2.17,14.85) | < 0.001 |
| Low C3 | 39 (81.2%) | 48 (43.6%) | 5.5 (2.44,13.11) | < 0.001 |
| Low C4 | 33 (68.8%) | 39 (35.5%) | 4.0 (1.85,8.46) | < 0.001 |
| Low complement C3/C4 | 39 (81.2%) | 56 (50.9%) | 4.1 (1.82,9.77) | < 0.001 |
| Coombs | 12 (25%) | 13 (11.8%) | 2.5 (0.96,6.24) | 0.056 |
| Elevated ESR | 37 (77.1%) | 82 (74.5%) | 1.2 (0.49,2.73) | 0.842 |
| ***At time of visit*** | | | | |
| SELENA-SLEDAI, mean (SD) | 2.5 (2,4) | 1 (0,2) | NA | <0.001 |
| PGA, mean (SD) | 0.5 (0,1) | 0.5 (0, 0.5) | NA | 0.537 |

| | | | | |
|---|---|---|---|---|
| C3, mean (SD) | 110.02 (36.95) | 135.63 (35.75) | NA | <0.001 |
| C4, mean (SD) | 18 (12, 27) | 24.5 (20, 32) | NA | <0.001 |
| Anti-dsDNA, mean (SD) | 10 (0, 80) | 0 (0,0) | NA | <0.001 |
| **SLEDAI (At time of visit)** | | | | |
| CNS | 0(0%) | 1 (0.9%) | NA | 1.000 |
| Vascular | 1 (2.2%) | 2 (1.9%) | 1.2 (0.10,12.98) | 1.000 |
| Renal | 3 (6.5%) | 5 (4.7%) | 1.4 (0.32,6.11) | 0.698 |
| Musculoskeletal | 4 (8.7%) | 5 (4.7%) | 1.9 (0.49,7.45) | 0.454 |
| Immunology | 27 (58.7%) | 16 (15.0%) | 7.6 (3.47,16.45) | <0.001 |
| Skin | 17 (37.0%) | 32 (29.9%) | 1.3 (0.65,2.75) | 0.451 |
| Serositis | 1 (2.2%) | 2 (1.9%) | 1.2 (0.10,13.27) | 1.000 |
| Hematology | 2 (4.4%) | 1 (0.9%) | 4.7 (0.41,53.6) | 0.215 |
| Constitutional | 0 (0%) | 0 (0%) | NA | NA |

OR : Odds ratio. CI: confidence interval. ESR: Erythrocyte sedimentation rate. SELENA-SLEDAI: Safety of Estrogens in Lupus National Assessment study-SLE disease activity index. PGA: physician global assessment. CNS: central nervous system. Comparisons were done using Fisher's exact test.
